# Social cognition after epilepsy surgery in temporal lobe epilepsy: A long-term follow-up

**DOI:** 10.64898/2025.12.06.25341481

**Authors:** Martin Simcik, Ross Andel, Viktoria Pytelova, Alena Javůrková, Jakub Žalud, Adam Kalina, Michaela Kalinova, Katerina Stanzelova, Petr Marusic, Jana Amlerova

## Abstract

**Background:** Previous studies have suggested that social cognition abilities do not change following surgical treatment for temporal lobe epilepsy (TLE). However, the follow-up period in these studies was no longer than 14 months. The present study investigated the long-term effects of epilepsy surgery on social cognition, extending the follow-up period to an average of 12 years (range 7-15 years).

**Methods:** We assessed 24 patients with drug-resistant TLE (mean age = 37 ± 11 years; 14 males) who underwent temporal lobe resection. Social cognition was measured using the Emotion Recognition Test (ERT, measuring emotion recognition) and the Faux-Pas Test (FPT, measuring Theory of Mind). Linear mixed-effects models accounted for repeated measures. Baseline scores for both ERT and FPT were standardized as Z-scores.

**Results:** ERT scores remained stable over time (estimate = 0.01, p = .405), while FPT performance declined gradually (estimate = −0.04, p = .009). Later epilepsy onset predicted higher initial FPT scores (estimate = 0.02, p = .036) but faster decline (estimate = −0.01, p = .013). Epilepsy duration, side, IQ, sex, and depressive symptoms had no significant effect.

**Conclusion:** While ERT appeared to remain stable over the extended period, FPT showed a gradual decline across the post-surgery follow-up independent of age. Later age at epilepsy onset was associated with steeper FPT deterioration. Our results extend previous findings, confirming stability in emotion recognition and suggesting decline in recognition of Faux-Pax after epilepsy surgery.

## Introduction

Social cognition, one of the six core domains of cognitive functioning, encompasses the mental processes involved in perceiving, interpreting, and responding to social stimuli^1^. It includes a range of functions, from basic abilities like recognizing emotions through facial expressions (facial emotion recognition, FER) to complex behavior such as being able to infer other people’s thoughts, feelings, and intentions (Theory of Mind, ToM)^2^. These processes are interconnected, and their integration is crucial for effective interpersonal communication^2^, because the inability to recognize facial emotions, such as, for example, a mournful face, can lead to impaired ToM and, in turn, worse well-being and lower quality of life (QoL)^3,4^. In other words, in social environments, individuals rely on intact social cognition to lead harmonious lives and maintain a satisfactory QoL.

The biological basis for these processes is viewed as a set of interconnected neural networks^5^, with their main hubs situated in the temporal and frontal lobes – specifically amygdala, anterior insula and ventrolateral prefrontal cortex for FER^6^, and ventromedial prefrontal cortex with temporoparietal junction for ToM^7^, and superior temporal sulcus and fusiform gyrus for both^7,8^. These distinct neural networks do not function in isolation. Instead, social cognition emerges from their coordinated and dynamic interactions, integrating perceptual and cognitive processes^5^. It has been shown that in TLE, the network that normally supports the recognition of fearful faces can collapse, with the amygdala failing to engage its usual partnered hubs during the task^9^. This interconnected nature of the brain suggests that pathologies – whether localized lesions or more widely distributed disruptions of neural networks^10^ – can impair social cognitive functioning. There is evidence that social cognition is impaired in many diseases affecting the temporal and frontal cortex, e.g. schizophrenia, Alzheimer disease or temporal lobe epilepsy (TLE)^11,12^.

In people with epilepsy, social cognition deficits significantly influence QoL, even when seizure freedom is achieved^4^. TLE patients show a high rate of pharmacoresistance and therefore they are often referred as candidates for epilepsy surgery. Surgical removal of an epileptogenic tissue offers the potential for full remission or at least for significant seizure reduction^13^. While seizure control is the primary goal, the effects of surgery on cognitive and psychosocial outcomes warrant closer examination. Standard neuropsychological domains, such as memory, executive function, and attention, are typically assessed one year after epilepsy surgery. A previous study by Amlerova et al. extended this research to include assessments of ToM and FER immediately before and 12 months after the surgery^14^. Neither significant improvements nor deterioration was found in social cognition at the group level, but several factors were related to poorer neuropsychological outcomes including longer epilepsy duration, earlier disease onset, and lower IQ. Despite these findings, the long-term impact of temporal lobectomy on social cognition remains an open question, highlighting the need for extended follow-up studies.

The aim of our study was to build upon the previous research by conducting an extended longitudinal follow-up of the same patients for up to 15 years (11.6 ± 2.2 years) to examine potential long-term post-surgical changes in social cognition. We also sought to assess whether any changes in FPT and ERT, if observed, would be modified by age of epilepsy onset, epilepsy duration, side of epilepsy, IQ performance, sex or depressive symptoms.

## Materials & Methods

### Participants

We followed 24 patients with TLE who had been previously investigated before temporal lobe resection for drug-resistant epilepsy at our epilepsy center. These individuals participated in a longitudinal study designed to assess social cognition impairments at three key timepoints: (1) prior to surgery, (2) 12 months after, and (3) 7 – 15 years after surgery. All participants had a confirmed diagnosis of unilateral mesial TLE based on electroclinical findings. Comprehensive preoperative assessments were conducted, incorporating neurological history and examination, MRI, routine and long-term video-EEG monitoring, and tests for language lateralization using either bilateral carotid sodium amobarbital/methohexital testing or fMRI. To ensure consistency, patients with atypical (bilateral or right hemisphere) speech dominance, significant cognitive impairment (Full-Scale IQ below 65), or seizures within 24 hours before testing were excluded from the study. All participants provided informed consent, and the study received approval from the institutional ethics committee.

### Measures

#### Social Cognition

To evaluate social cognition abilities, we administered the Emotion Recognition Test (ERT) to assess FER and Faux Pas Test (FPT) to measure ToM. The ERT, adapted from the Ekman and Friesen series^15^, consisted of 25 photographs showcasing facial expressions of five emotions: happiness, fear, sadness, disgust, and anger. We omitted the surprise expression prior to testing, due to its frequent confusion with fear^16^, as including this highly ambiguous category could decrease the sensitivity of the overall score. Participants were tasked with matching each facial expression to one of the emotion labels listed beneath the photo. There was no time limit for responses. The maximum score for ERT was 25 points. The FPT, a concise version adapted from Schacher et al.^10^, involved reading three brief stories, each featuring a social faux pas. With the text available in front of them, participants answered questions to detect the faux-pas and deduce the characters’ mental states. Although we followed Schacher et al. in selecting only three stories, we applied the full six-question scoring structure from the original adult FPT developed by Baron-Cohen and colleagues^17^, as used in previous work from our center^14^. A correct response required both to rightly assess a social faux pas, and to understand the characters’ mental state. If a patient interpreted the situation as intentionally harmful, the answers were marked as incorrect. If no faux pas was identified, two control questions were presented to assess the text comprehension, though these did not contribute to the final score. The maximum FPT score was 18, and like the ERT, there was no time restriction for responses.

#### Covariates

Demographic factors included age (in years) and sex (male/female). Clinical factors encompassed age at epilepsy onset, duration of epilepsy, epilepsy laterality (left/right), and International League Against Epilepsy (ILAE)^18^ outcome. For the ILAE outcome, only the assessment of the last year from the long-term follow-up was considered. Neuropsychological assessments included Full-Scale IQ (FSIQ), measured using the Wechsler Adult Intelligence Scale – Third Edition (WAIS-III)^19^ and depressive symptoms, evaluated with the Beck Depression Inventory (BDI)^20^.

### Analyses

Statistical analyses were performed in R, version 4.4.2^21^, with the *lme4* package for linear mixed-effects models^22^ and the *lmerTest* package for obtaining p-values and model diagnostics^23^. To account for the repeated measures design (with three measurement occasions), we used mixed-effects models^24^, treating time as a fixed effect and including random intercepts for individual patients. Mixed effects models are an advanced and popular approach to modeling longitudinal data^25^. They can effectively account for individual variability in scores by modeling random effects, contain multiple options for covariance matrix specification, and contain other advantages that provide overall better fit between the data and the specified model. FPT and ERT scores were converted to Z-scores (mean = 0, SD = 1) using the baseline (T1) distribution of the patient sample. Consequently, values at T2 and T3 represent standardized deviations from the cohort’s pre-operative performance. Covariates were centered to the sample mean for better model interpretability.

Initially, we calculated intraclass correlations for each outcome using unconditional models with only the random intercepts and no fixed effects. In order to establish the main analytical model, we a) tested covariance structure in a model with time as the sole independent variable and b) sequentially tested additional covariates, quadratic effects of time, and explored potential interactions to determine whether they improved model fit. Model comparisons were conducted using the likelihood ratio test (p > .05), which is the primary method designated for this purpose in R. To determine ideal covariance structure, we compared model fit of the compound symmetry, unstructured, and autoregressive structures. While all structures produced similar fixed effects results, the unstructured matrix provided better fit than compound symmetry (χ2= 109.9, p < .001) and autoregressive (χ2= 109.7, p < .001).

Using this covariance structure, we observed that covariates age, sex, side of epilepsy, epilepsy duration, and depressive symptoms did not improve the fit of neither FPT nor ERT model. Therefore, to create a parsimonious model and to be mindful of presumed bias towards Type II error due to the small sample size, the final model treating FPT as the outcome included time, age of epilepsy onset, FSIQ, and an interaction between time and onset. For ERT, the final model included time and FSIQ as fixed effects, with a random intercept for individual patients. The main analyses were estimated with restricted maximum likelihood because this approach provides less biased variance estimates than other estimation methods including the full maximum likelihood method, especially when sample size is small^26^.

### Handling Age of Epilepsy Onset

We explored categorizing patients into Early (n= 12) and Late onset (n= 12) groups using a cutoff at 20 years of age when considering ToM. This threshold aligns partially with recent research on ToM development, which suggests that FPT performance does not stabilize until young adulthood^25^. It is important to note that this categorization may reflect general cognitive maturation rather than a fixed developmental endpoint for FPT^27^. However, when we compared models, treating age of onset as a continuous variable consistently outperformed the binary Early/Late split in terms of model fit. Therefore, we opted to keep age of onset as a continuous variable in the final FPT model.

We did not categorize onset age for ERT, as emotion recognition develops earlier than ToM, with core skills emerging by age 5 and refining by age 12^28^. Prior studies on emotion recognition in TLE used a 5-year cutoff to distinguish early and late onset^29^, but applying this threshold in our sample would have resulted in an Early onset group too small for analysis (n = 3). Therefore, we automatically retained the onset age as a continuous variable.

Finally, we consider previous research indicating that people with epilepsy can follow different trajectories of cognitive change depending on several clinical and psychosocial factors^30^. Hence, we also explored individual changes that could be obscured by group-level results, as well as patterns using previously published criterion, which defines a significant change in ERT or FPT as a shift of three or more points between two timepoints^14^.

## Results

### Demographic information

In our sample of 24 patients, males (n = 14) outnumbered females (n = 10), and left-sided TLE (n = 15) was slightly more common than right-sided TLE (n = 9). The mean age at baseline was 37.2 ± 10.5 years. The mean age of epilepsy onset 19.7 ± 12.2 years, range 1 – 47 years. In the last year of the long-term follow-up, based on the ILAE postoperative seizure outcome classification, 20 of the 24 patients were classified as outcome 1, one patient had an outcome of 3, and three were classified as ILAE outcome 4. An overview of baseline characteristics is provided in *Table 1*.

**Table 1.** Baseline Characteristics.

|  | **All Patients (n =24)** | **LTLE (n = 15)** | **RTLE (n = 9)** |
| --- | --- | --- | --- |
| **Mean Age (SD), years** | 37.2 ± 10.5 | 34.4 ± 10.8 | 42.0 ± 8.5 |
| **Age Range, years** | 21–57 | 21–55 | 28–57 |
| **Sex (M/F)** | 14.10 | 8.7 | 6.3 |
| **Mean Age at onset (SD)** | 19.7 ± 12.4 | 19.1 ± 10.7 | 20.7 ± 15.6 |
| **Age of onset Range, years** | 1–47 | 3–44 | 1–47 |
| **Mean epilepsy duration (SD), years** | 17.8 ± 11.2 | 15.4 ± 8.4 | 21.7 ± 14.5 |
| **Mean ERT (SD)** | 20.7 ± 3.6 | 20.5 ± 3.9 | 21.0 ± 3.2 |
| **Range of ERT scores** | 11–25 | 11–25 | 14–25 |
| **Mean FPT (SD)** | 12.9 ± 4.8 | 13.5 ± 3.8 | 11.8 ± 6.2 |
| **Range of FPT scores** | 0–18 | 4–18 | 0–18 |
| **Mean FSIQ (SD)** | 99.7 ± 15.4 | 98.6 ± 13.6 | 101.6 ± 18.7 |
| **Mean BDI (SD)** | 9.1 ± 7.6 | 12.0 ± 7.4 | 4.1 ± 5.1 |
| **Mean follow-up period (SD), years** | 11.6 ± 2.2 | 11.6 ± 2.2 | 11.6 ± 2.4 |
| **Range of follow-up period, years** | 7–15 | 8–15 | 7–15 |
| *Abbreviations: BDI, Beck Depression Inventory; ERT, emotion recognition test; FPT, faux-pas test; FSIQ, full-scale IQ; F, females; M, males; LTLE, left temporal lobe epilepsy; RTLE, right temporal epilepsy; SD, standard deviation.* |  |  |  |

### Unconditional models

The intraclass correlation for FPT was 32%, indicating that approximately one-third of its variance stemmed from differences between patients, while the remaining 68% reflected variance in scores over time (i.e., within patients). These results suggest that FPT scores tended to change within individuals over time more than between patients at baseline. In contrast, ERT’s intraclass correlation was 58%, indicating that more than half of the variance was between patients with the remaining 42% indicating variability of scores within patients over time. In other words, ERT scores appeared slightly more stable over time than FPT scores but still showed observable variability. Performance scores on the ERT and FPT at each of the three time points are presented in *Table 2*.

**Table 2.** Cognitive performance across the three measurement occasions.

| **Cognition tests** | **Side of epilepsy** | **Pre-surgery** | **12 Months after surgery** | **~ 11.6 Years after surgery** |
| --- | --- | --- | --- | --- |
| **ERT** | *mean (SD)* |  |  |  |
|  | Both sides | 20.7 (3.6) | 20.7 (3.8) | 21 (3) |
|  | LTLE | 20.5 (3.9) | 21.3 (2.4) | 21.2 (2.2) |
|  | RTLE | 21.0 (3.2) | 19.7 (5.3) | 20.7 (4.1) |
|  | *range* |  |  |  |
|  | Both sides | 11–25 | 11–25 | 14–25 |
|  | LTLE | 11–25 | 18–24 | 17–24 |
|  | RTLE | 14–25 | 11–25 | 14–25 |
| **FPT** | *mean (SD)* |  |  |  |
|  | Both sides | 12.9 (4.8) | 12.1 (4.3) | 10 (3.6) |
|  | LTLE | 13.5 (3.8) | 12.5 (3.8) | 10.5 (3.9) |
|  | RTLE | 11.8 (6.2) | 11.4 (5.2) | 9.2 (2.9) |
|  | *range* |  |  |  |
|  | Both sides | 0–18 | 2–17 | 4–17 |
|  | LTLE | 4–18 | 4–17 | 4–17 |
|  | RTLE | 0–18 | 2–17 | 4–12 |
| *Abbreviations: ERT, emotion recognition test, FPT, faux-pas test; LTLE, left temporal lobe epilepsy; RTLE; right temporal lobe epilepsy; SD, standard deviation.* |  |  |  |  |

### FPT as the outcome

Results from the mixed-effects models are presented in *Table 3*. There was a positive relationship between baseline FPT score and age of epilepsy onset (estimate = 0.02, p = .036), alongside a general declining trend in FPT scores over time (estimate = −0.04, p = .009). This base rate of FPT decline was greater in individuals with later epilepsy onset (estimate = −0.003, p = .013). Furthermore, FSIQ showed a positive association with FPT at the baseline (estimate = 0.03, p < .001). Considering that the original model included both seizure free patients (n = 20) and patients with ILAE outcome > 1 (n = 4), we conducted a post-hoc analysis focusing on seizure-free patients only. This approach yielded similar results for the effect of time (estimate = −0.04, p = .013), and the interaction between the effect of time and age of epilepsy onset (estimate = −0.003, p = .012).

**Table 3.** Results of the main mixed effects models.

| Parameter | Estimate | CI |  | p-value |
| --- | --- | --- | --- | --- |
|  |  | CI low | CI high |  |
| ERT model |  |  |  |  |
| Fixed effects |  |  |  |  |
| Intercept | -0.05 | -0.38 | 0.27 | .739 |
| Time | 0.01 | -0.02 | 0.04 | .405 |
| FSIQ (centered) | 0.03 | 0.01 | 0.04 | .005 |
| Random effects |  |  |  |  |
| Variance (intercept) | 0.59 |  |  |  |
| Variance (residual) | 0.68 |  |  |  |
| FPT model |  |  |  |  |
| Fixed effects |  |  |  |  |
| Intercept | 0.17 | -0.09 | 0.43 | .198 |
| Time | -0.04 | -0.07 | -0.01 | .009 |
| FSIQ (centered) | 0.04 | 0.02 | 0.05 | <.001 |
| Age of onset (centered) | 0.02 | 0.00 | 0.05 | .036 |
| Interaction between Time and Onset | -0.01 | -0.01 | -0.00 | .013 |
| Random effects |  |  |  |  |
| Variance (intercept) | 0.37 |  |  |  |
| Variance (residual) | 0.70 |  |  |  |
| Abbreviations: CI, confidence intervals; ERT, emotion recognition test; FPT, faux-pas test; FSIQ, full-scale IQ |  |  |  |  |

### ERT as the outcome

Overall, ERT scores improved slightly and non-significantly across the study period (estimate = 0.01, p = .405). In contrast, FSIQ showed a significant positive association with ERT scores at the baseline (estimate = 0.02, p = .005) but did not otherwise interact with subsequent postoperative changes.

### Individual Changes

While group level analyses showed no overall change in the case of ERT, and a gradual decline in FPT, individual trajectories displayed considerable variation (*Figures 1, 2*). These trajectories suggest that assuming a linear change may not fully capture the underlying patterns. Instead, they reveal considerable fluctuation, with many patients (14/24 for ERT, 13/24 for FPT) showing both initial improvement and subsequent decline (5/24 for ERT, 9/24 for FPT), and initial decline and subsequent improvement (9/24 for ERT, 4/24 for FPT). To assess long-term changes based on previously published criteria^13^, we compared the first and last timepoints of the study using this criterion. This analysis revealed both notable improvements (4/24 for ERT, 4/24 for FPT) and declines (4/24 for ERT, 11/24 for FPT), where the remaining patients showed no significant change (16/24 for ERT, 9/24 for FPT).

**Figure 1:**
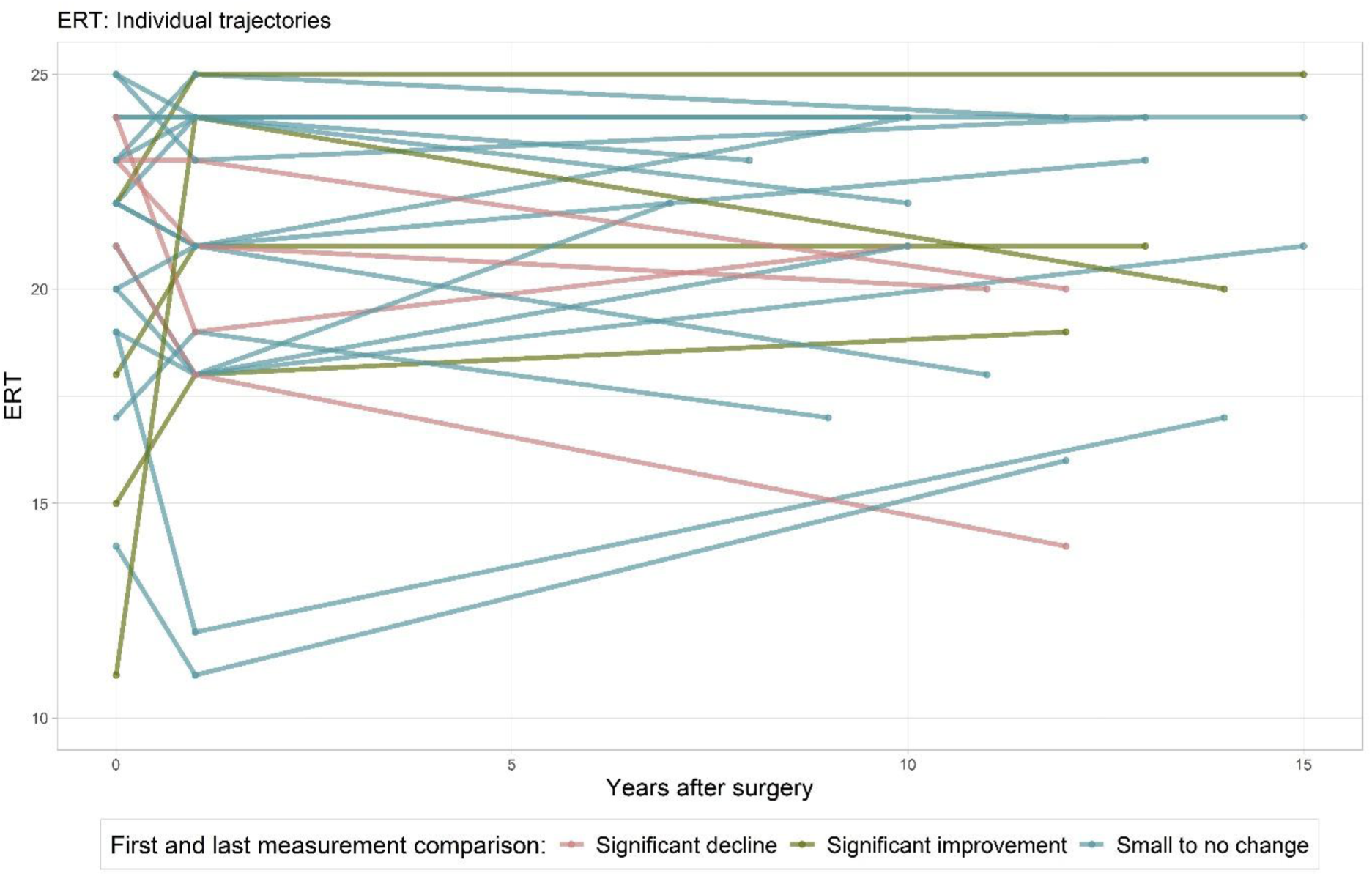
Emotion Recogntion Test (ERT) – Individual trajectories. Spaghetti plot showing ERT scores of 24 patients across three time points. Each line represents an individual patient. Color indicates the magnitude and direction of change between the first and last measurement: green lines represent patients with a significant improvement (≥3-point increase), red lines indicate a significant decline (≥3-point decrease), and blue lines represent non-significant change (change <3 points). Significant change (either direction) is defined as a minimum difference of 3 points between the first and last time point.

**Figure 2:**
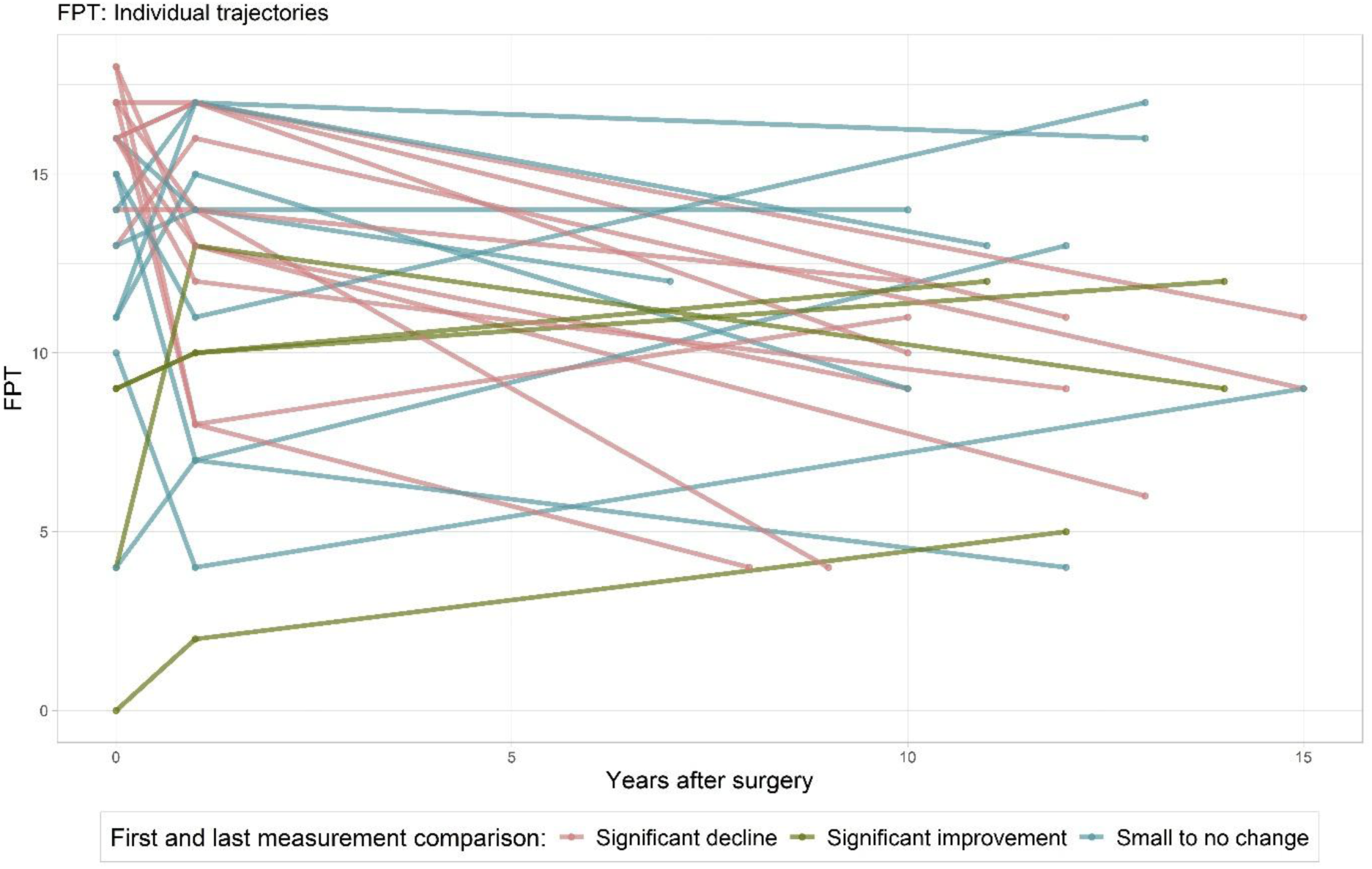
Faux-pas Test (FPT) – Individual trajectories. Spaghetti plot showing FPT scores of 24 patients across three time points. Each line represents an individual patient. Color indicates the magnitude and direction of change between the first and last measurement: green lines represent patients with a significant improvement (≥3-point increase), red lines indicate a significant decline (≥3-point decrease), and blue lines represent non-significant change (change <3 points). Significant change (either direction) is defined as a minimum difference of 3 points between the first and last time point.

## Discussion

We investigated the long-term trajectory of social cognition in patients with TLE following epilepsy surgery, extending the follow-up period from 1 year used in Amlerova et al. (2014) to an average of 11.6 ± 2.2 years after surgery. Our findings indicate that, on a group level, ERT remained stable over time whereas FPT performance decreased gradually after surgery in a generally linear fashion. Overall later age of epilepsy onset was associated with higher baseline performance, but also with a faster rate of decline on the FPT. Furthermore, as expected, higher FSIQ scores were associated with better baseline performance on both tests. Notably, neither ERT nor FPT were significantly influenced by sex, epilepsy duration, side of epilepsy, or depressive symptoms at baseline or in terms of change in scores over time. Together, our results indicate that social cognition after epilepsy surgery exhibits long-term stability, with potential decline in recognition of faux pas overtime.

To the best of our knowledge, our study is the first to track indicators of social cognition trajectories in long term period post-surgery, offering valuable insights into its evolution after anteromedial temporal lobe resection. In a recent meta-analysis^31^, it has been stated that neither emotion recognition nor ToM abilities change after epilepsy surgery. However, individual longitudinal studies have had typically limited follow-up durations, often being capped at one year^14,32–35^.

When examining individual patterns more closely, the subgroup with clinically meaningful change (≥3 points) in either ERT, FPT, or both (N = 18) showed that substantial improvement across both measures was uncommon, where only a single patient with left-sided TLE demonstrated this pattern. At the opposite end, only one individual exhibited a major decline on both tasks, in this case with right-sided TLE. Notably, both outlier cases had an ILAE outcome of 1 and experienced their first seizures in early childhood (age range 0-5 and 6-10, respectively), suggesting that factors shaping these long-term trajectories reach beyond early onset and long-term postoperative seizure freedom. Major declines in FPT (−10 to −13 points; N = 3) occurred alongside stable ERT performance, reinforcing the idea that ToM and ER rely on partly independent mechanisms. All three pronounced FPT decliners had left-sided TLE and later epilepsy onset (age range 26-30, and 40-45). Finally, all patients with an ILAE outcome greater than 1 (N = 4) were present within this subgroup, and those with an ILAE outcome of 4 did not show significant improvement on either test.

A brief exploratory review of clinical stability according to ILAE outcomes between the 12-month and long-term follow-up showed that postoperative seizure outcome remained unchanged in 22 of 24 patients. Only two individuals experienced worsening in ILAE outcome (patient #6: 1→3; patient #15: 3→4). Their social-cognitive trajectories did not reveal any pattern, where would seizure recurrence lead to change in the test scores: patient #6 showed improvement in FPT (9→10→12) and decline in ERT (23→21→20), whereas patient #15 scored consistently higher and remained relatively stable across both measures (FPT 16→14→14; ERT 24→19→21). Post-operative changes in ASM load were similarly sparse, with only two patients increasing the number of medications. Patient #15 (ASM count 3→4) remained broadly stable in social cognition, as mentioned above, while patient #17 (ASM count 2→3) improved on the FPT (+6 points) and declined slightly on the ERT (–3 points). These different patterns of FPT and ERT change did not point to any consistent link with either ASM count or ILAE outcome. Because seizure outcomes and ASM adjustments showed very little variation in our sample, the data are too limited to support firm conclusions about their impact on long-term social-cognitive performance.

In terms of lateralization, we did not find any effect of the epilepsy side on the outcome variables, which aligns with a prior short-term study on the same sample^14^, although we concede that the small sample size in the side-specific analyses may have obscured the results. Two prior studies found mixed results concerning the relationship between emotion recognition and side following an epilepsy surgery; Shaw (2007)^32^ found an improvement in processing fearful faces only in left-sided TLE, but Benuzzi (2014)^33^ discovered an enhanced fear recognition performance in two patients with early onset right-sided TLE. Furthermore, postoperative improvements in alternative measures of ToM, such as performance on false-belief tasks, were noted, even when FPT scores remained unchanged^34^. These discrepancies underline the limitations of relying on only two relatively simple tests, which may not fully encompass the multifaceted nature of social cognition.

Broader evidence not limited to post-surgical differences suggests that right-sided TLE patients perform worse than left-sided TLE patients in social cognition, particularly in ToM tasks^36^. Our data aligns with this notion, showing lower pre-surgical FPT means in right-sided TLE patients, though postoperative FPT changes appear similar across both sides (*Table 2*). ToM relies on distributed fronto-temporo-parietal networks^5^, and these can be disrupted by chronic epilepsy^37^. The pre-surgical deficits in right-sided TLE then likely reflect greater interference with right-lateralized components of this bilateral system. Resective surgery, by eliminating seizure foci and ceasing associated network disruption, may enable partial functional recovery in both groups. Importantly, the longitudinal data available to this date (although limited to 14 months after surgery) show that ToM performance tends to remain stable post-operatively^31^, suggesting that resective surgery does not worsen social cognition and may even allow partial recovery by removing seizure-driven network disruption. In an extreme demonstration of neural plasticity, a recent case series reported that four individuals who underwent hemispherectomy between 3 months and 20 years of age exhibited substantial unilateral reorganization of ToM-related responses within the remaining hemisphere^38^. These observations imply that neither hemisphere is uniquely indispensable for supporting ToM processing and that the right hemisphere is not rigidly required for socioemotional function. However, broader claims about typical lateralization or the general principles of network reorganization remain outside the scope of current evidence.

Taken together, these findings highlight that the core mechanisms underlying social cognition impairment in epilepsy are likely multifactorial. Structural changes, such as epileptic lesions in the temporal poles, which are richly interconnected hubs processing and storing social and emotional information^39^, alongside interictal epileptic brain activity^40^ and the notion that epilepsy can be characterized as a network disorder, disrupting connections and impairing the functionality of distant brain regions, ultimately affecting multiple cognitive domains^41–44^. Based on these considerations, it has been hypothesized that surgical removal of a portion of the richly interconnected temporal lobe could either further compromise social cognitive functioning by removing key neural substrates or, conversely, facilitate network reorganization that may lead to cognitive improvement in seizure-free patients^32,45^. Moreover, the social stigma associated with epilepsy and the severity of seizures may contribute to social isolation, thereby limiting opportunities for social engagement^46^. Finally, the sudden transition to a seizure-free life can present its own challenges, potentially giving rise to new psychosocial issues^47^. Therefore, a reciprocal influence between this complex postoperative adjustment process and the restoration of social cognitive abilities may arise.

It has been pointed out that group comparisons can hide individual post operative dynamics^31^, and our findings support this view. Looking at individual patients, a considerable proportion displayed non-linear, complex trends in their social cognitive trajectories, where initial improvements in either FPT or ERT scores were followed by subsequent declines at later follow-up points, and vice versa. These disparate early postoperative changes could reflect individual differences in the pace of neural and cognitive adaptation. Assessing every patient at a fixed interval (12 months) may therefore capture transient worsening for some and ongoing improvement for others, yielding heterogeneous trends that only later converge toward stability on the group level. The limited number of assessments, only three timepoints, may have been insufficient to capture the true trajectory of change. Furthermore, unaccounted factors such as the degree of social reintegration post-surgery could also contribute to these unexpected patterns. Ultimately, following patients after epilepsy surgery should consider clinical, psychological, as well as social aspects^48^.

Our results should be interpreted with caution. The uneven follow-up periods present a significant methodological challenge, as patients were reassessed at disparate intervals, potentially confounding the interpretation of longitudinal trends. Moreover, the small sample size may have yielded limited statistical power to detect subtle effects of clinical variables, such as the side of surgery and demographic factors, which could mean that small, yet clinically relevant influences remain undetected. The inclusion of both seizure-free patients and those with less favorable ILAE outcomes may have also influenced the observed patterns in cognitive change. Finally, we did not take into account anti-seizure medication. Although its effects on cognition could have influenced the outcomes^49^, the potential confounding effect should not be too large, as other cognitive abilities, in our case FSIQ, did not show a significant change across the years.

Interpreting long-term change without a control group also requires caution. Normal ageing involves some decline in social cognition that is partly independent of general cognitive decline^50^. However, if ageing alone were driving change in our cohort, one would expect a broadly consistent pattern across individuals. This was not the case, as the trajectories in both ERT and FPT were markedly heterogeneous. This pronounced inter-individual variability argues against a simple ageing explanation and supports the idea that long-term postoperative outcomes in social cognition are shaped by factors beyond natural ageing alone. Nonetheless, in the absence of a non-surgical TLE group or healthy control group, we cannot definitively quantify how much of the observed variance reflects disease-specific, surgery-related, or age-related influences. Future studies with appropriately matched controls are needed to disentangle these contributions.

The choice of paradigms also warrants consideration. In ERT, we relied on static black-and-white facial photographs, which may underestimate subtle deficits, as dynamic stimuli have shown greater sensitivity in TLE^51^. However, because the baseline assessment was conducted 15 years ago, we retained the Ekman and Friesen Pictures of Facial Affect to ensure comparability across the 11.8-year follow-up interval. This limitation should be considered when interpreting the magnitude of change. In contrast, the FPT, though also an established instrument, continues to serve as a sensitive and clinically relevant measure of Theory of Mind in neurological populations, including TLE^51^.

A further limitation is that the pronounced intraindividual variability and the small sample size limit the stability of slope estimates and increase both Type I and Type II error risks, even when using mixed-effects models^52^. Consequently, the observed FPT change should be interpreted cautiously, and replication in larger cohorts with more frequent follow-up assessments is needed.

Finally, mixed-effects models were selected because they account for within-person correlation and efficiently handle repeated-measures data, even in small samples, without discarding incomplete cases. Evidence indicates that linear mixed models produce unbiased fixed effect estimates in modest samples when multiple observations per participant are available^53,54^. A post-hoc Monte-Carlo power analysis based on our study design suggested adequate power (71-99%) to detect medium-to-large effects. Nevertheless, given the sample size, results should be interpreted cautiously and replicated in larger cohorts.

## Conclusion

Our study suggests that the postoperative dynamics of emotion recognition and ToM differ following epilepsy surgery. While the ability to recognize others’ emotions appears to remain largely stable at the group level, the capacity to infer others’ mental states declines gradually over time, particularly in patients with later epilepsy onset who initially exhibit higher performance. The presence of both notable improvements and declines among individual patients suggests that the small sample size may have masked more subtle effects, rendering them statistically non-significant. However, it is unlikely that large effects would emerge with the inclusion of additional patients. At the same time, the limited sample size increases the possibility that some of the observed changes, particularly the FPT decline, could represent false-positive findings shaped by unmeasured factors rather than true postoperative effects Alternatively, it is also possible that additional factors, such as social integration or occupational status, may be influencing the outcomes. Nevertheless, the small sample size also may have caused a false effect altogether, as countless other unmeasured factors may have influenced these results. Future research should test this possibility, preferably with larger samples and more homogeneity with respect to clinical outcome and laterality of the temporal lobe affected. More frequent and evenly spaced follow-up assessments may also be preferable to capture the temporal dynamics of social cognition following epilepsy surgery.

## Acknowledgments

This work was supported by IPE No 699012 and NPO, the project National Institute for Neurological Research (Programme EXCELES, ID Project No. LX22NPO5107) – Funded by the European Union – Next Generation EU. The work was also supported by the Czech Alzheimer Foundation and Epilepsy Research Centre in Prague (EpiReC). Author Micheala Kalinova was supported by ERDF-Project Brain dynamics, No. CZ.02.01.01/00/22_008/0004643.

## Ethics approval statement

We confirm that we have read the Journal’s position on issues involved in ethical publication and affirm that this report is consistent with those guidelines.

## Data availability statement

The dataset supporting the conclusions of this article is available in the Open Science Framework repository [DOI 10.17605/OSF.IO/2ZBH4] and can also be accessed via Zenodo [DOI 10.5281/zenodo.15602632].

## References

1. During the preparation of this work the author(s) used Elicit in order to effectively search for relevant literature. After using this tool/service, the author(s) reviewed and edited the content as needed and take(s) full responsibility for the content of the published article.

1. Adolphs, R. The neurobiology of social cognition. Curr. Opin. Neurobiol. 11, 231–239 (2001).

2. Mitchell, R. L. C. & Phillips, L. H. The overlapping relationship between emotion perception and theory of mind. Neuropsychologia 70, 1–10 (2015).

3. Marafioti, G., Cardile, D., Culicetto, L., Quartarone, A. & Lo Buono, V. The Impact of Social Cognition Deficits on Quality of Life in Multiple Sclerosis: A Scoping Review. Brain Sci. 2024 Vol 14 Page 691 14, 691–691 (2024).

4. Yogarajah, M. & Mula, M. Social cognition, psychiatric comorbidities, and quality of life in adults with epilepsy. Epilepsy Behav. EB 100, (2019).

5. Krendl, A. C. & Betzel, R. F. Social cognitive network neuroscience. Soc. Cogn. Affect. Neurosci. 17, 510–529 (2022).

6. Xu, P., Peng, S., Luo, Y. jia & Gong, G. Facial expression recognition: A meta-analytic review of theoretical models and neuroimaging evidence. Neurosci. Biobehav. Rev. 127, 820–836 (2021).

7. Molenberghs, P., Johnson, H., Henry, J. D. & Mattingley, J. B. Understanding the minds of others: A neuroimaging meta-analysis. Neurosci. Biobehav. Rev. 65, 276–291 (2016).

8. Skiba, R. M. & Vuilleumier, P. Brain Networks Processing Temporal Information in Dynamic Facial Expressions. Cereb. Cortex 30, 6021–6038 (2020).

9. Broicher, S. D. et al. Alterations in functional connectivity of the amygdala in unilateral mesial temporal lobe epilepsy. J. Neurol. 259, 2546–2554 (2012).

10. Schacher, M. et al. Mesial temporal lobe epilepsy impairs advanced social cognition. Epilepsia 47, 2141–2146 (2006).

11. Amlerova, J. et al. Emotional prosody recognition is impaired in Alzheimer’s disease. Alzheimers Res. Ther. 14, (2022).

12. Duclos, H., Desgranges, B., Eustache, F. & Laisney, M. Impairment of social cognition in neurological diseases. Rev. Neurol. (Paris) 174, 190–198 (2018).

13. Sperling, M., O’Connor, M., Saykin, A. & Plummer, C. Temporal lobectomy for refractory epilepsy. JAMA 276, 470–470 (1996).

14. Amlerova, J. et al. Emotion recognition and social cognition in temporal lobe epilepsy and the effect of epilepsy surgery. Epilepsy Behav. 36, 86–89 (2014).

15. Ekman, P. & Friesen, W. V. Pictures of facial affect. Consult. Psychol. Press (1976).

16. Rapcsak, S. Z. et al. Fear recognition deficits after focal brain damage: A cautionary note. Neurology 54, 575–581 (2000).

17. Stone, V. E., Baron-Cohen, S. & Knight, R. T. Frontal Lobe Contributions to Theory of Mind. J. Cogn. Neurosci. 10, 640–656 (1998).

18. Wieser, H. G. et al. Proposal for a New Classification of Outcome with Respect to Epileptic Seizures Following Epilepsy Surgery. Epilepsia 42, 282–286 (2001).

19. Wechsler, D. Wechsler Adult Intelligence Scale. 3rd Edition. Psychol. Corp. San Antonio (1997).

20. Beck, A. T. Beck Depression Inventory–II (BDI-II). (1996).

21. R Core Team. R: A Language and Environment for Statistical Computing. (2024).

22. Bates D., Machler M., Bolker, B. & Walker, S. Fitting Linear Mixed-Effects Models Using lme4. 67, 1–48 (2015).

23. Kuznetsova, A., Brockhoff, P. B. & Christensen, R. H. B. lmerTest Package: Tests in Linear Mixed Effects Models. J. Stat. Softw. 82, 1–26 (2017).

24. Yu, Z. et al. Beyond t-Test and ANOVA: applications of mixed-effects models for more rigorous statistical analysis in neuroscience research. Neuron 110, 21–21 (2021).

25. Verbeke, G., Molenberghs, G. & Rizopoulos, D. Random Effects Models for Longitudinal Data. in Longitudinal Research with Latent Variables (eds van Montfort, K., Oud, J. H. L. & Satorra, A.) 37–96 (Springer, Berlin, Heidelberg, 2010). doi:10.1007/978-3-642-11760-2_2.

26. McNeish, D. Multilevel Mediation With Small Samples: A Cautionary Note on the Multilevel Structural Equation Modeling Framework. Struct. Equ. Model. Multidiscip. J. 24, 609–625 (2017).

27. Meinhardt-Injac, B., Daum, M. M. & Meinhardt, G. Theory of mind development from adolescence to adulthood: Testing the two-component model. Br. J. Dev. Psychol. 38, 289–303 (2020).

28. Stewart, E. et al. Facial emotion perception and social competence in children (8 to 16 years old) with genetic generalized epilepsy and temporal lobe epilepsy. Epilepsy Behav. 100, 106301–106301 (2019).

29. Meletti, S. et al. Impaired facial emotion recognition in early-onset right mesial temporal lobe epilepsy. Neurology 60, 426–431 (2003).

30. Baxendale, S. The who, what, when and why of cognitive difficulties in epilepsy. Br. J. Hosp. Med. 84, 1–8 (2023).

31. Mikula, B., Lencsés, A., Borbély, C. & Demeter, G. Emotion recognition and theory of mind after temporal lobe epilepsy surgery: A systematic review. Seizure 93, 63–74 (2021).

32. Shaw, P. et al. A prospective study of the effects of anterior temporal lobectomy on emotion recognition and theory of mind. Neuropsychologia 45, 2783–2790 (2007).

33. Benuzzi, F. et al. Recovery from emotion recognition impairment after temporal lobectomy. Front. Neurol. 5 JUN, 92037–92037 (2014).

34. Giovagnoli, A. R., Parente, A., Didato, G., Deleo, F. & Villani, F. Expanding the spectrum of cognitive outcomes after temporal lobe epilepsy surgery: A prospective study of theory of mind. Epilepsia 57, 920–930 (2016).

35. Grangé, M. et al. Facial emotion recognition in focal epilepsy: localization is not the main factor. Epilepsy Behav. 172, (2025).

36. Ziaei, M., Arnold, C., Thompson, K. & Reutens, D. C. Social Cognition in Temporal and Frontal Lobe Epilepsy: Systematic Review, Meta-analysis, and Clinical Recommendations. J. Int. Neuropsychol. Soc. JINS 29, 205–229 (2023).

37. van Diessen, E., Diederen, S. J. H., Braun, K. P. J., Jansen, F. E. & Stam, C. J. Functional and structural brain networks in epilepsy: what have we learned? Epilepsia 54, 1855–1865 (2013).

38. Kliemann, D., Adolphs, R., Paul, L. K., Tyszka, J. M. & Tranel, D. Reorganization of the Social Brain in Individuals with Only One Intact Cerebral Hemisphere. Brain Sci. 11, 965 (2021).

39. Olson, I. R., Plotzker, A. & Ezzyat, Y. The Enigmatic temporal pole: a review of findings on social and emotional processing. Brain 130, 1718–1731 (2007).

40. Laufs, H. et al. Temporal lobe interictal epileptic discharges affect cerebral activity in “default mode” brain regions. Hum. Brain Mapp. 28, 1023–1032 (2007).

41. Bernhardt, B. C., Hong, S., Bernasconi, A. & Bernasconi, N. Imaging structural and functional brain networks in temporal lobe epilepsy. Front. Hum. Neurosci. 7, 50812–50812 (2013).

42. Viskontas, I. V., McAndrews, M. P. & Moscovitch, M. Remote Episodic Memory Deficits in Patients with Unilateral Temporal Lobe Epilepsy and Excisions. J. Neurosci. 20, 5853–5857 (2000).

43. Bartha-Doering, L. & Trinka, E. The interictal language profile in adult epilepsy. Epilepsia 55, 1512–1525 (2014).

44. Hwang, G. et al. Cognitive slowing and its underlying neurobiology in temporal lobe epilepsy. Cortex 117, 41–52 (2019).

45. Edwards, M., Stewart, E., Palermo, R. & Lah, S. Facial emotion perception in patients with epilepsy: A systematic review with meta-analysis. Neurosci. Biobehav. Rev. 83, 212–225 (2017).

46. Steiger, B. K. & Jokeit, H. Why epilepsy challenges social life. Seizure 44, 194–198 (2017).

47. Wilson, S. J., Bladin, P. F. & Saling, M. M. The burden of normality: A framework for rehabilitation after epilepsy surgery. Epilepsia 48, 13–16 (2007).

48. Kirsch, H. E. Social cognition and epilepsy surgery. Epilepsy Behav. 8, 71–80 (2006).

49. Eddy, C. M., Rickards, H. E. & Cavanna, A. E. The cognitive impact of antiepileptic drugs. Ther. Adv. Neurol. Disord. 4, 385–407 (2011).

50. Kemp, J., Després, O., Sellal, F. & Dufour, A. Theory of Mind in normal ageing and neurodegenerative pathologies. Ageing Res. Rev. 11, 199–219 (2012).

51. Bauer, J. et al. A comparative study of social cognition in epilepsy, brain injury, and Parkinson’s disease. PsyCh J. 12, 443–451 (2023).

52. Schielzeth, H. & Forstmeier, W. Conclusions beyond support: overconfident estimates in mixed models. Behav. Ecol. 20, 416–420 (2009).

53. Wiley, R. W. & Rapp, B. Statistical analysis in Small-N Designs: using linear mixed-effects modeling for evaluating intervention effectiveness. Aphasiology 33, 1–30 (2019).

54. Serrano, D. Error of estimation and sample size in the linear mixed model. in (2008).

